# Risk factors for interstitial lung disease in early rheumatoid arthritis and external validation of screening strategies: Baseline results of SAIL-RA

**DOI:** 10.1101/2025.01.24.25321091

**Authors:** Gregory C McDermott, Ritu Gill, Suzanne Byrne, Staci Gagne, Xiaosong Wang, Misti L Paudel, Emily Kowalski, Grace Qian, Katarina Bade, Kevin Mueller, Alene Saavedra, Kathleen MM Vanni, Liya Sisay Getachew, Caleb Bolden, Lauren A O’Keeffe, Natalie A Davis, Alison Puri, Tina Mahajan, Erica Mulcaire-Jones, Neda Kortam, Pierre-Antoine Juge, Tracy J Doyle, Paul F Dellaripa, Zachary S Wallace, Raul San Jose Estepar, George R Washko, Marcy B Bolster, Kevin D. Deane, Dinesh Khanna, Bryant R England, Jeffrey A Sparks

## Abstract

**Background:** Risk factors and screening strategies for rheumatoid arthritis-associated interstitial lung disease (RA-ILD) have received limited evaluation in patients with early RA. We investigated RA-ILD prevalence, risk factors, and the performance of proposed RA-ILD screening methodologies in a multicenter, prospective study of patients with early RA.

**Methods:** Participants with early RA, defined as being within two years of RA diagnosis, were enrolled at five US sites and assessed with high-resolution computed tomography (HRCT) chest imaging, pulmonary function tests, and autoantibodies. RA-ILD presence was determined through independent HRCT review by thoracic radiologists. We investigated RA-ILD risk factors using multivariable logistic regression and reported the predictive performance of RA-ILD screening strategies (ANCHOR-RA, 2023 ACR/CHEST, Four Factor Score, and ESPOIR).

**Results:** Among 172 participants (74% female, 82% seropositive, median RA duration 0.79 years, mean age 55.3 years), 19 (11%) had ILD on HRCT. Moderate/high RA disease activity by DAS28-ESR (OR 7.00 [1.95, 25.1]) and age ≥60 years (OR 3.87 [1.33, 11.3]) were associated with RA-ILD. Sensitivity and specificity of screening strategies ranged from 0.32-0.95 and 0.32-0.81, respectively. The number of early RA patients needing screening to detect one ILD case ranged from 3.6 to 6.4.

**Discussion:** In this prospective, multicenter study, ILD prevalence in early RA was 11%. Disease activity and older age were strongly associated with ILD in early RA, and several proposed ILD screening strategies performed showed promise for enabling ILD screening in early RA.

**KEY MESSAGES:** *What is already known on this topic:*

- Several risk factors for rheumatoid arthritis-associated interstitial lung disease have been identified. However, most prior studies have focused on patients with established RA.
- Several approaches to RA-ILD screening have been proposed, including the recent 2023 ACR/CHEST guidelines. However, none have been specifically evaluated in patients with early RA.

*What this study adds:*

- In a multicenter, prospective cohort of patients with early RA (diagnosed within 2 years of enrollment), moderate or high RA disease activity and older age were significantly associated with evidence of interstitial lung disease on high resolution CT chest imaging.
- Several proposed RA-ILD screening criteria performed well in the early RA period. The simplest screening strategy included older age, male sex, and moderate/high RA disease activity and had a number needed to screen of 3.6 patients to detect one RA-ILD case.

*How this study might affect research, practice or policy:*

- Simple screening strategies using demographic and clinical data may enable selection of early RA patients for RA-ILD screening.

## INTRODUCTION

Rheumatoid arthritis-associated interstitial lung disease (RA-ILD) is a severe, extra-articular disease manifestation of RA that is associated with increased morbidity, mortality, and health care costs[1–8]. There has been increasing recognition that RA-ILD may occur throughout the RA disease course[9], including prior to the onset of articular disease[2,10–13] and shortly after diagnosis in the “early RA” period[14–19]. Despite this recognition, there have been limited investigations performing systematic screening for RA-ILD in patients with early RA. To better understand the prevalence and risk factors for RA-ILD and other RA-related lung diseases in early RA, we performed a multicenter prospective screening study of RA patients with recently diagnosed RA.

Previous estimates of the prevalence of both subclinical and clinically-apparent RA-ILD vary widely based on the specific case criteria and screening methods used as well as the population studied. Given the plausible mechanisms linking mucosal inflammation in sites like the lung to the pathogenesis and development of RA[20], as well as the possibility that early RA may present a “window of opportunity” where effective disease-modifying treatment may alter articular progression[21–23], studying lung disease in patients with newly diagnosed RA is of particular interest. However, there has been limited research systematically screening for lung disease in a recently diagnosed RA population.

Several RA-ILD screening strategies have been proposed based on known epidemiologic risk factors for lung disease in RA[24]. The recently published 2023 ACR/CHEST guidelines for screening and monitoring of ILD in patients with systemic autoimmune diseases conditionally recommended screening asymptomatic patients with autoimmune diseases, including RA, who are at increased risk of lung disease with autoimmune diseases, including RA[25]. Risk factors for ILD among RA patients include high titer rheumatoid factor (RF) and anti-cyclic citrullinated peptide (anti-CCP) antibodies, cigarette smoking, older age at RA onset, high disease activity, male sex, and higher body mass index[25]. A recent investigation led by the ESPOIR investigators derived and independently validated a risk score for subclinical RA-ILD, which included male sex, older age, RA disease activity, and the *MUC5B* promoter variant, a known genetic risk factor for RA-ILD[26,27]. Other recent studies have suggested weighted risk scores that include combinations of demographic and lifestyle risk factors along with autoantibodies and/or inflammatory marker testing[28–30]. Finally, the ongoing multinational ANCHOR-RA study, which is investigating RA-ILD prevalence in higher risk populations selected five recognized epidemiologic risk factors as inclusion criteria[31]. However, the performance of many of these screening strategies in independent populations, especially in patients with early RA, is unclear.

To investigate the prevalence and risk factors for RA-ILD and other RA-related lung diseases in the early RA period, we performed a multicenter, prospective study of patients with early RA. All patients were assessed with comprehensive lung screening including HRCT chest imaging, pulmonary function tests, laboratory testing, and RA disease activity measures. We hypothesized that RA disease activity would be strongly associated with the presence of ILD in the early RA period. Subsequently, we aimed to evaluate the performance of previously proposed RA-ILD screening strategies through external validation in this early RA population.

## METHODS

### Study design and population

We performed a cross-sectional analysis of the ongoing longitudinal prospective Study of Inflammatory Arthritis and Interstitial Lung Disease in Early RA (SAIL-RA). Study participants were recruited between 2017 and 2024 at five academic healthcare centers in the United States: Brigham and Women’s Hospital (Boston, MA), Massachusetts General Hospital (Boston, MA), University of Nebraska Medical Center (Omaha, NE), University of Colorado (Aurora, CO), and University of Michigan (Ann Arbor, MI). All participants were diagnosed with RA within 2 years prior to study enrollment and met 2010 ACR/EULAR RA classification criteria[32]. These study participants are designated herein as “early RA.”

The study was approved by the Mass General Brigham (#2020P003768) and University of Nebraska Medical Center (#0282-16-FB) Institutional Review Boards. Written informed consent was obtained from all participants. Patients and the public were not involved in the development of the study protocol.

### Study measures

At the baseline study visit, all patients were assessed with surveys, history, physical examination, high-resolution CT (HRCT) chest imaging, pulmonary function tests, and laboratory testing. Survey data included demographics including age, sex, self-reported race, and ethnicity, respiratory exposures including smoking status (never, past, or current smoker), pack-years of smoking, and the Modified Medical Research Council dyspnea scale[33]. HRCT scans were performed on multidetector CT scanners (at least 16 detectors) with inspiratory, supine volumetric images and image reconstruction using 1 to 1.25mm slice thickness. Pulmonary function testing included forced expiratory volume in one second (FEV_1_), forced vital capacity (FVC), and diffusion capacity of the lungs for carbon monoxide (DLCO). Laboratory testing for erythrocyte sedimentation rate (ESR) was performed at baseline. Data on rheumatoid factor, and anti-cyclic citrullinated peptide was obtained from review of clinically performed labs. Physical examination included assessment of height and weight, which was used to calculate body mass index (BMI). Cardiopulmonary examination and tender and swollen joint assessments were performed by trained rheumatologists at all sites.

Disease Activity Score with 28 joints and ESR (DAS28-ESR) was calculated for each patient based on tender and swollen joint counts, laboratory testing, and patient global assessment obtained at the study visit[34,35]. The disease activity was further characterized into remission or low (DAS28-ESR <3.2) or moderate or high (DAS28-ESR ≥3.2). In 12 patients who were missing ESR, the Clinical Disease Activity Index[35] was used to determine disease activity category. Current medications, RA symptom duration, and RA diagnosis date were obtained from review of electronic health records.

### HRCT interpretation

Each research HRCT was independently reviewed by at least two thoracic radiologists. Each scan was scored by two thoracic radiologists for the presence of interstitial lung abnormalities using established criteria that defined interstitial lung abnormalities as nondependent changes affecting >5% of any lobar area[36] as well as the presence or absence of bronchiectasis and emphysema. Discordant scoring was resolved by a third radiologist who independently reviewed the scan and whose score was used as a tiebreaker.

For each patient with evidence of ILD, one thoracic radiologist performed visual scoring to determine semiquantitative extent and pattern of ILD involvement. The scoring system was based the CT chest scoring used in the Scleroderma Lung Study I[37,38]. Briefly, each scan was divided into three lung zones (upper, middle, lower) and scored for the presence and extent of ground glass opacities, fibrosis, honeycombing, emphysema in each zone. Subtypes were determined using established criteria for non-usual interstitial pneumonia (UIP), typical/definite UIP, probable UIP, or indeterminate for UIP[38] as well as established guidelines for typical non-UIP radiologic patterns (e.g., hypersensitivity pneumonitis)[39,40].

### Previous RA-ILD Screening Strategies

We assessed the eligibility criteria for the ongoing ANCHOR-RA screening study for RA-ILD[24,31], risk factors suggested in the 2023 ACR/CHEST ILD screening/monitoring guidelines[25], the risk factors identified in our study, which were overlapping with those proposed from the ESPOIR cohorts[26], and a recently proposed weighted “Four Factor” risk score[28]. The ANCHOR-RA eligibility criteria included two or more of: high-titer RF and/or anti-CCP autoantibodies (>3x ULN), presence of extra-articular RA disease manifestations (nodules, Sjögren’s syndrome, or vasculitis), ever smoking status, male sex, RA onset at or after age 60, and moderate or high RA disease activity. The ACR/CHEST factors included high-titer RF and/or anti-CCP antibodies, ever smoking status, moderate or high RA disease activity, and obesity. The ESPOIR risk factors, which coincided with those identified in our multivariable models, included male sex, older age of RA onset, and moderate/high disease activity. Of note, the ESPOIR study also considered the *MUC5B* promoter variant in an extended model, but this was not available in SAIL-RA or in routine clinical care[26]. Finally, the Four Factor weighted risk score included age at RA onset, smoking status, and RF and anti-CCP autoantibody titers in the score[28].

### Statistical analysis

We reported descriptive statistics for the cohort including mean and standard deviation (SD) or median and interquartile range (IQR) for continuous variables. We constructed unadjusted logistic regression models examining the association between age, sex, smoking status, RF and anti-CCP autoantibody status, and RA disease activity with presence of ILD and reported the results as odds ratios (OR) and 95% confidence intervals (CI). These models categorized age at RA diagnosis as greater than or equal to 60 years, consistent with prior research on RA-related risk factors[41,42] and also categorized RF and anti-CCP titers into negative, low positive (>1 to 3x the upper limit of normal [ULN]), and high positive (>3x ULN) according to the reference range of the performing laboratory. Based on statistically significant results from our unadjusted models, we created a multivariable model that adjusted for age, sex, and RA disease activity. For these models, the comparator group was RA participants without any RA-related lung disease, excluding a small number of participants who had emphysema or bronchiectasis without ILD were excluded.

In secondary analyses, we repeated the analysis examining the outcome of any parenchymal lung disease (ILD, emphysema, or bronchiectasis) compared to RA without lung disease. In unadjusted models, RF was associated with smoking status and any parenchymal lung disease for this outcome, so the multivariable model additionally adjusted for smoking status and categorical RF level along with age, sex, and disease activity. We also performed a sensitivity analysis that included these additional factors in the models investigating ILD as the outcome. Finally, to examine possible sex differences, we stratified by sex at birth. We created a multivariable model adjusted for age and RA disease activity based on the results of the unadjusted models after sex stratification.

Finally, we examined the performance of suggested RA-ILD screening strategies for identifying ILD cases in our cohort. For each set of criteria, we examined the sensitivity, specificity, positive predictive value (PPV), and negative predictive value (NPV) along with 95%CIs calculated using the Clopper-Pearson method for those with low cell sizes[43]. ANCHOR-RA enrollment required two or more risk factors and the Four Factor Score used a 5-point cutoff, while other strategies either suggested consideration of screening among patients with one or more risk factor or did not specify a strict cutoff. Thus, we compared the sensitivity, specificity, PPV, and NPV of each criteria at different cutoffs. First, we examined the presence of one or more factors for ACR/CHEST, ANCHOR-RA, ESPOIR/SAIL-RA to investigate more sensitive screening approaches. We then examined the screening performance for the same models using two or more factors as well as the 5-point cutoff of the Four Factor Score to compare strategies with more specific screening approaches. We report number needed to screen (NNS) to detect one RA-ILD case for these screening strategies and calculated area under the receiver operating curves (AUROC) for each set of criteria.

As screening for respiratory symptoms may not be performed universally or in a structured fashion in rheumatology clinics, we chose to include all early RA participants in our validation of RA-ILD screening strategies. In a sensitivity analysis, we limited to RA patients who had a Modified Medical Research Council Dyspnea score less than 3[44].

A two-sided p-value of <0.05 was considered statistically significant. All analyses were performed using SAS version 9.4 (Cary, NC).

## RESULTS

### Study sample and baseline characteristics

We analyzed a total of 172 participants with early RA. Baseline characteristics of the cohort as well as subsets of participants with RA-ILD, bronchiectasis or emphysema without RA-ILD, and no evidence of any parenchymal lung disease are detailed in **Table 1**. The mean (SD) age of the participants was 55.3 years (13.7), 74.4% were female, 83.7% reported White race, and 61.6% were never smokers. Characteristics of the patients with any parenchymal lung disease (ILD, bronchiectasis, and/or emphysema) are detailed in **Supplementary Table S1**.

**Table 1:**
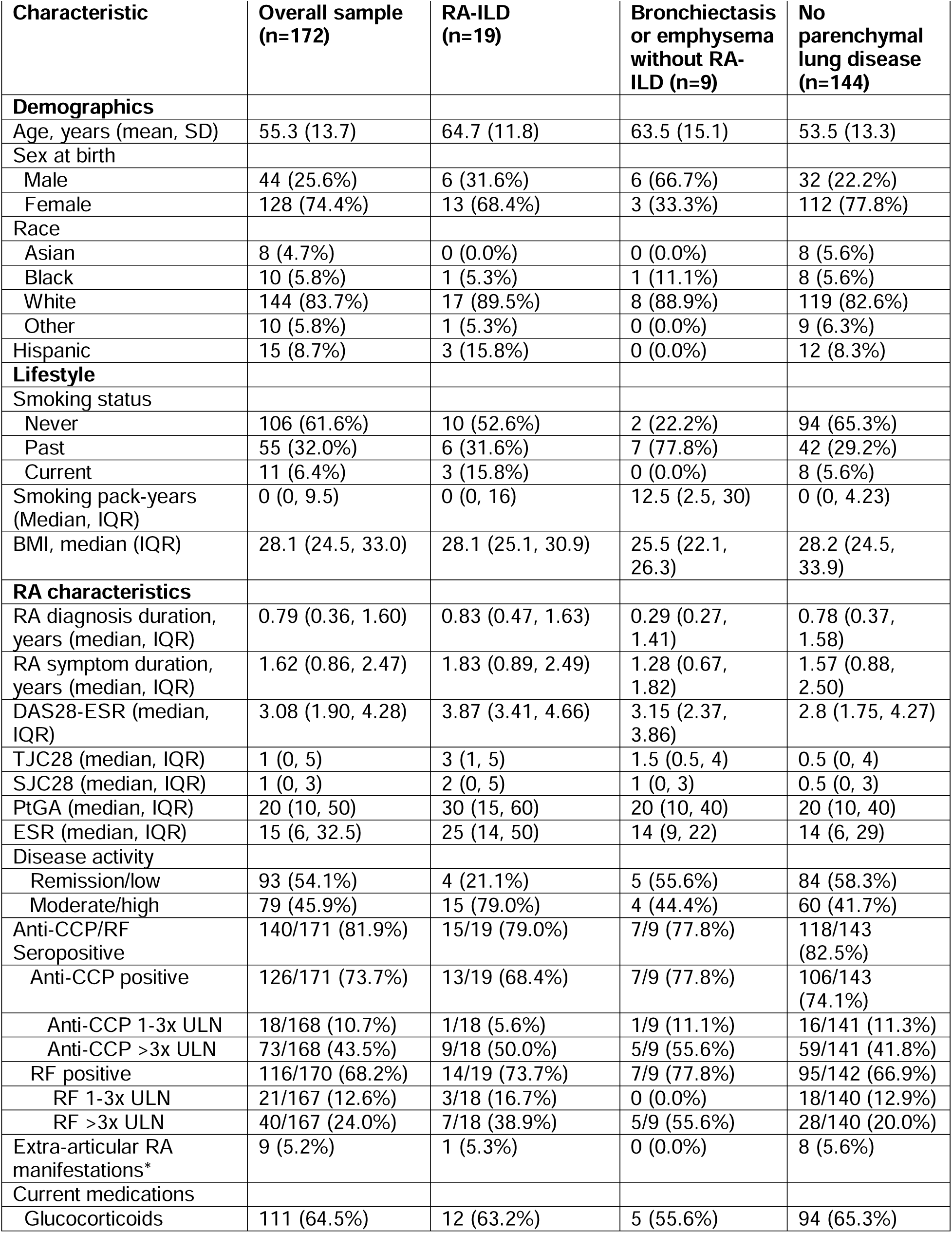
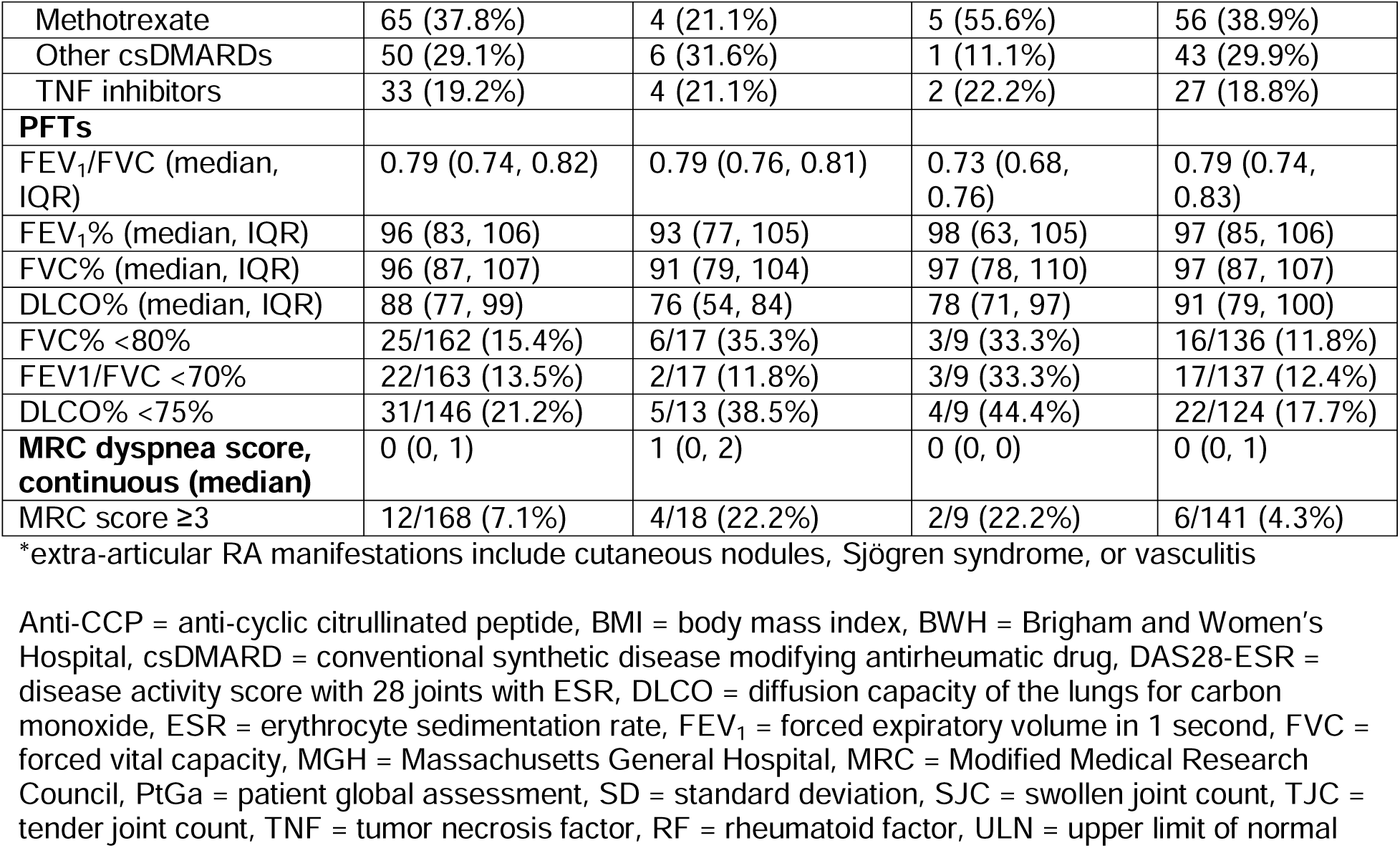
Baseline characteristics of SAIL-RA participants, overall and by presence or absence of ILD or other parenchymal lung disease on high-resolution computed tomography imaging.

The median RA disease duration at baseline was 0.79 years (IQR 0.36, 1.60). Seropositive participants accounted for 81.9% of the cohort, and 45.9% of the cohort was in moderate or high disease activity at the time of baseline visit, with 64.5% using glucocorticoids. *Prevalence of parenchymal lung disease*

The prevalence of parenchymal lung diseases is shown in **Figure 1**. RA-ILD was detected in a total of 19 participants (11.0%). Bronchiectasis was detected in 9 participants (5.2%), and emphysema in 7 participants (4.1%). Both RA-ILD and bronchiectasis overlap were noted in 5 participants (2.9%). The majority of participants (83.7%) did not have HRCT evidence of parenchymal lung disease.

**Figure 1:**
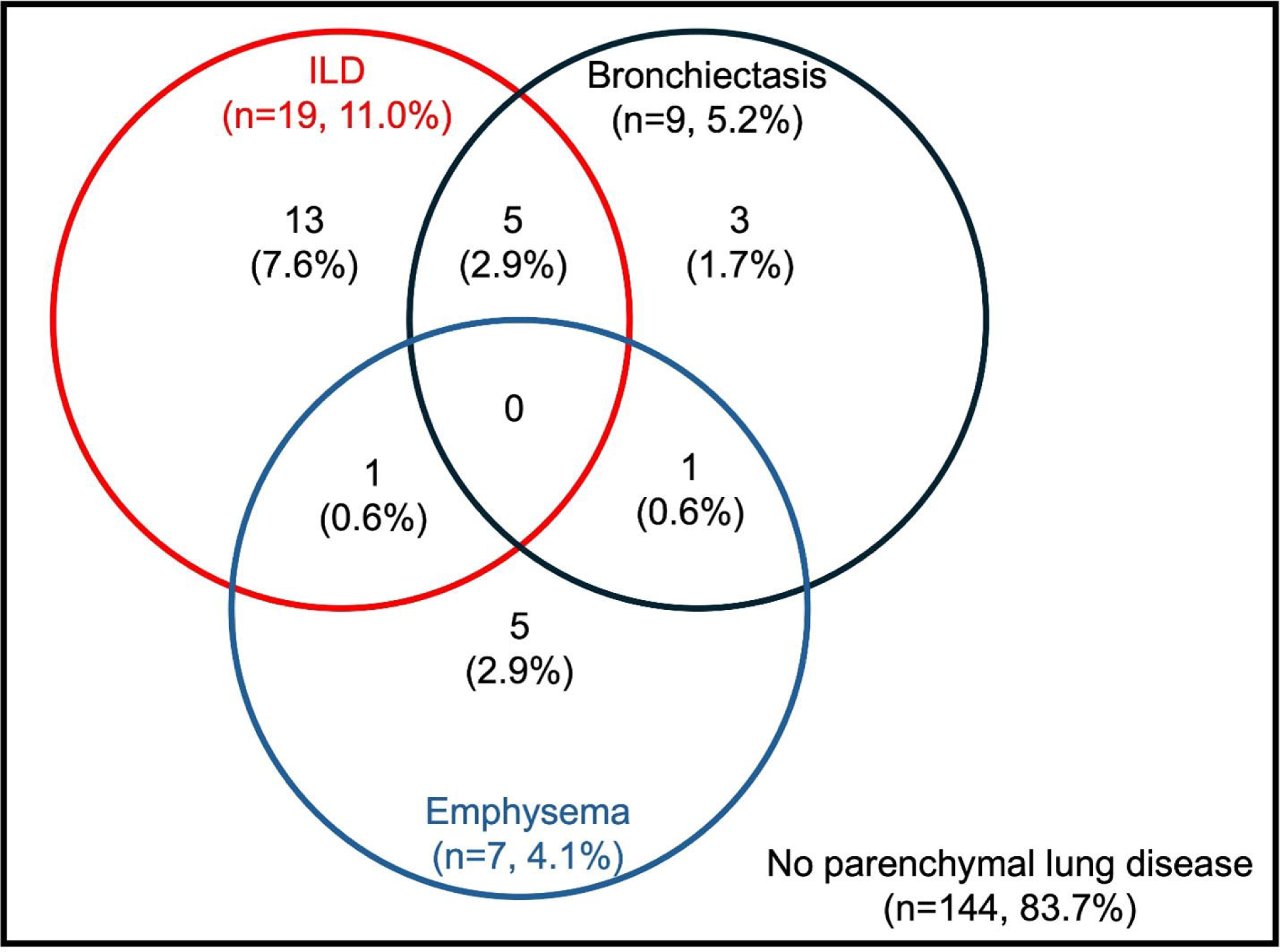
Overlap of interstitial lung disease, bronchiectasis, and emphysema in SAIL-RA, a prospective multicenter cohort of early RA patients (n=172) Presence of RA-related lung diseases was determined by review of high-resolution CT chest imaging of all participants by up to three expert thoracic radiologists. ILD = interstitial lung disease, RA = rheumatoid arthritis

### ILD severity, extent, and subtypes

Results of the semiquantitative and subtype scoring for the RA-ILD patients, as well as available PFT data, are detailed in **Supplemental Table S2**. The majority with RA-ILD (12/19, 63.2%) had 0-10% of lung involvement while 5/19 (26.3%) had 10-30% involvement and 2/19 (10.5%) patient had >30% involvement. A total of 7/19 RA-ILD patients (36.8%) did not have classifiable ILD subtype while 5/19 patients (26.3%) had nonspecific interstitial pneumonia pattern and 5/19 (26.3%) had UIP pattern. Only 6 participants with RA-ILD had FVC <80% predicted, including only 1 of 6 participants with >10% lung involvement by chest CT.

### Risk factors for ILD in early RA

Factors associated with RA-ILD are shown in **Table 2**. After adjustment, moderate/high disease activity had an OR for RA-ILD of 7.00 (95%CI 1.95 to 25.1) compared to remission/low disease activity. Age ≥60 years at RA diagnosis had an OR for RA-ILD of 3.87 (95%CI, 1.33 to 11.3) compared to <60 years. We saw similar results in the models that were additionally adjusted for RF level and smoking status (**Supplemental Table S3**). In analyses stratified by sex, estimates of the association between moderate/high disease activity and RA-ILD were strongest among female participants, with overlapping confidence intervals between male and female participants (**Supplemental Table S4**).

**Table 2:**
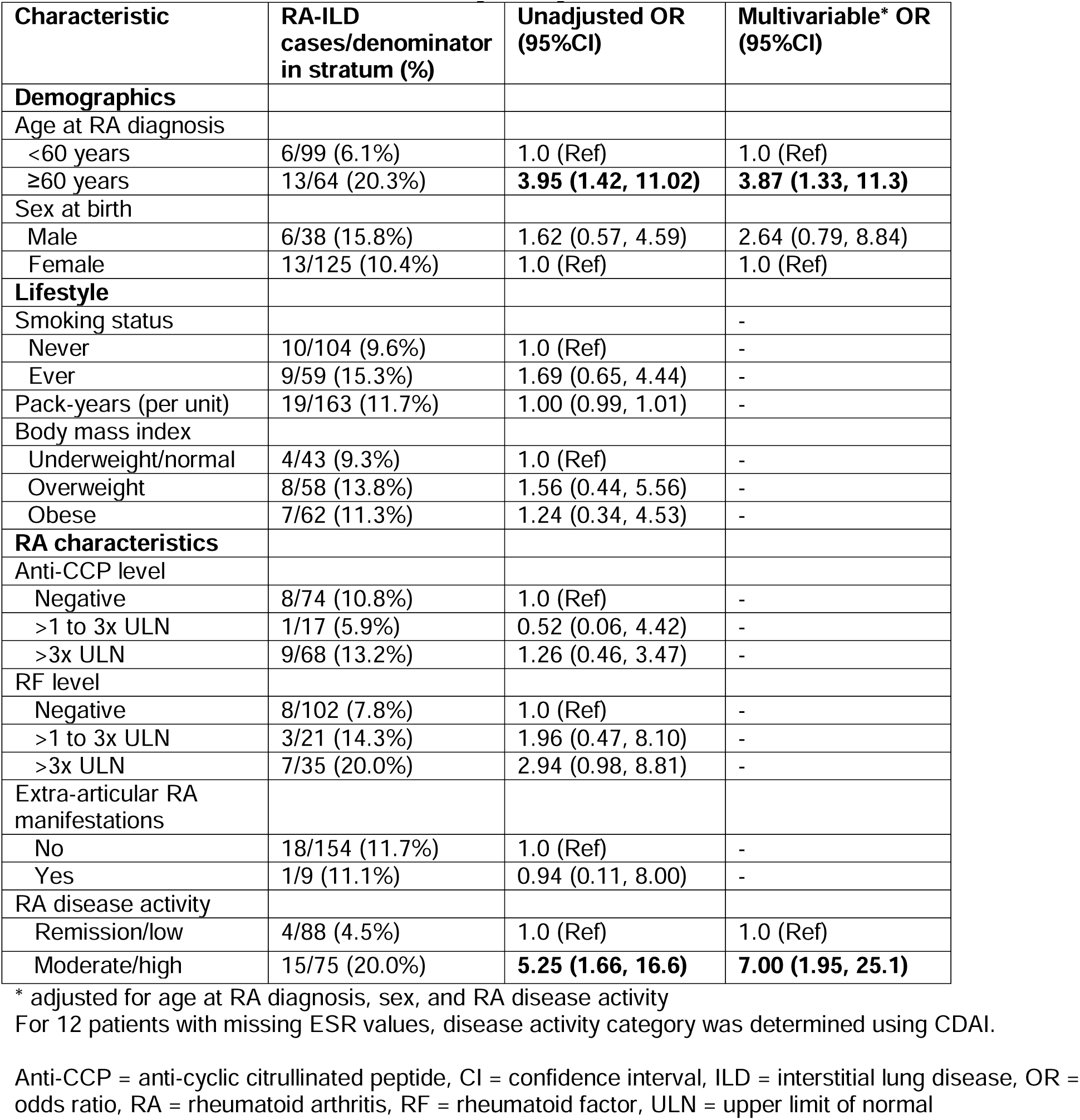
Odds ratios for RA-ILD in early RA by baseline characteristics.

### Risk factors for any parenchymal lung disease

Associations of baseline factors with any parenchymal lung disease are detailed in **Table 3**. Moderate/high disease activity (OR 3.60, 95%CI 1.24 to 10.5) was strongly associated with any parenchymal lung disease. Additional factors associated with any parenchymal lung disease were age ≥60 years (OR 3.61, 95%CI 1.38 to 9.46) and male sex (OR 3.47, 95%CI 1.23 to 9.83). RF and smoking status were no longer significant in the multivariable model.

**Table 3:** Odds ratios for any parenchymal lung disease (ILD, bronchiectasis, or emphysema)

| Characteristic | Parenchymal lung disease cases/denominator in stratum | Unadjusted OR (95%CI) | Multivariable* OR (95%CI) |
| --- | --- | --- | --- |
| **Demographics** |  |  |  |
| Age at RA diagnosis |  |  |  |
| <60 years | 8/101 (7.9%) | 1.0 (Ref) | 1.0 (Ref) |
| ≥60 years | 20/71 (28.2%) | **4.56 (1.88, 11.08)** | **3.61 (1.38, 9.46)** |
| Sex at birth |  |  |  |
| Male | 12/44 (27.3%) | **2.63 (1.13, 6.11)** | **3.47 (1.23, 9.83)** |
| Female | 16/128 (12.5%) | 1.0 (Ref) | 1.0 (Ref) |
| **Lifestyle** |  |  |  |
| Smoking status |  |  |  |
| Never | 12/106 (11.3%) | 1.0 (Ref) | 1.0 (Ref) |
| Ever | 16/66 (24.2%) | **2.51 (1.10, 5.71)** | 1.76 (0.69, 4.53) |
| Pack-years (per unit) | 28/172 (16.3%) | 1.00 (0.99, 1.01) | - |
| Body mass index |  |  |  |
| Underweight/normal | 8/47 (17.0%) | 1.0 (Ref) | - |
| Overweight | 11/61 (18.0%) | 1.07 (0.39, 2.92) | - |
| Obese | 9/64 (14.1%) | 0.80 (0.28, 2.25) | - |
| **RA characteristics** |  |  |  |
| Anti-CCP level |  |  |  |
| Negative | 11/77 (14.3%) | 1.0 (Ref) | - |
| >1 to 3x ULN | 2/18 (11.1%) | 0.75 (0.15, 3.72) | - |
| >3x ULN | 14/73 (19.2%) | 1.42 (0.60, 3.38) | - |
| RF level |  |  |  |
| Negative | 12/106 (11.3%) | 1.0 (Ref) | 1.0 (Ref) |
| >1 to 3x ULN | 3/21 (14.3%) | 1.31 (0.33, 5.10) | 1.13 (0.27, 4.80) |
| >3x ULN | 12/40 (30.0%) | **3.36 (1.36, 8.30)** | 2.16 (0.78, 5.98) |
| Extra-articular RA manifestations |  |  |  |
| No | 27/163 (16.6%) | 1.0 (Ref) | - |
| Yes | 1/9 (11.1%) | 0.63 (0.08, 5.24) | - |
| RA disease activity |  |  |  |
| Remission/low | 9/93 (9.7%) | 1.0 (Ref) | 1.0 (Ref) |
| Moderate/high | 19/79 (24.1%) | **2.96 (1.25, 6.98)** | **3.60 (1.24, 10.5)** |

\* adjusted for age at RA diagnosis, sex, smoking status, RF level, and RA disease activity For 12 patients with missing ESR values, disease activity category was determined using CDAI.
Anti-CCP = anti-cyclic citrullinated peptide, CI = confidence interval, ILD = interstitial lung disease, OR = odds ratio, RA = rheumatoid arthritis, RF = rheumatoid factor, ULN = upper limit of normal

### External validation of RA-ILD screening strategies

We assessed the performance of proposed screening strategies for RA-ILD in early RA (**Table 4**). When using a cut-off of at least one risk factor, the sensitivity was high for ANCHOR-RA, 2023 ACR/CHEST, and ESPOIR/SAIL-RA strategies (range 0.95 to 1.0) and the specificity was poor (range 0.11 to 0.30). When increasing the cut-off to at least two risk factors, the sensitivity was modestly reduced to 0.95 for both ANCHOR-RA and 2023 ACR/CHEST while specificity modestly improved to 0.40 for ANCHOR-RA and 0.32 for 2023 ACR/CHEST strategies. With this increased cut-off, the ESPOIR/SAIL-RA strategy had a sensitivity of 0.79 and specificity of 0.73, resulting in the highest PPV of 0.28. When using a 5-point cutoff of the Four Factor Score, the sensitivity and specificity were 0.32 and 0.81, respectively. The ESPOIR/SAIL-RA criteria had the highest OR per factor (OR per factor 3.97, 95%CI 1.88 to 8.40), followed by ANCHOR-RA (OR per factor 2.20, 95%CI 1.39 to 3.48), and ACR/CHEST criteria (OR per factor 1.85, 95%CI 1.25 to 2.73).

**Table 4:**
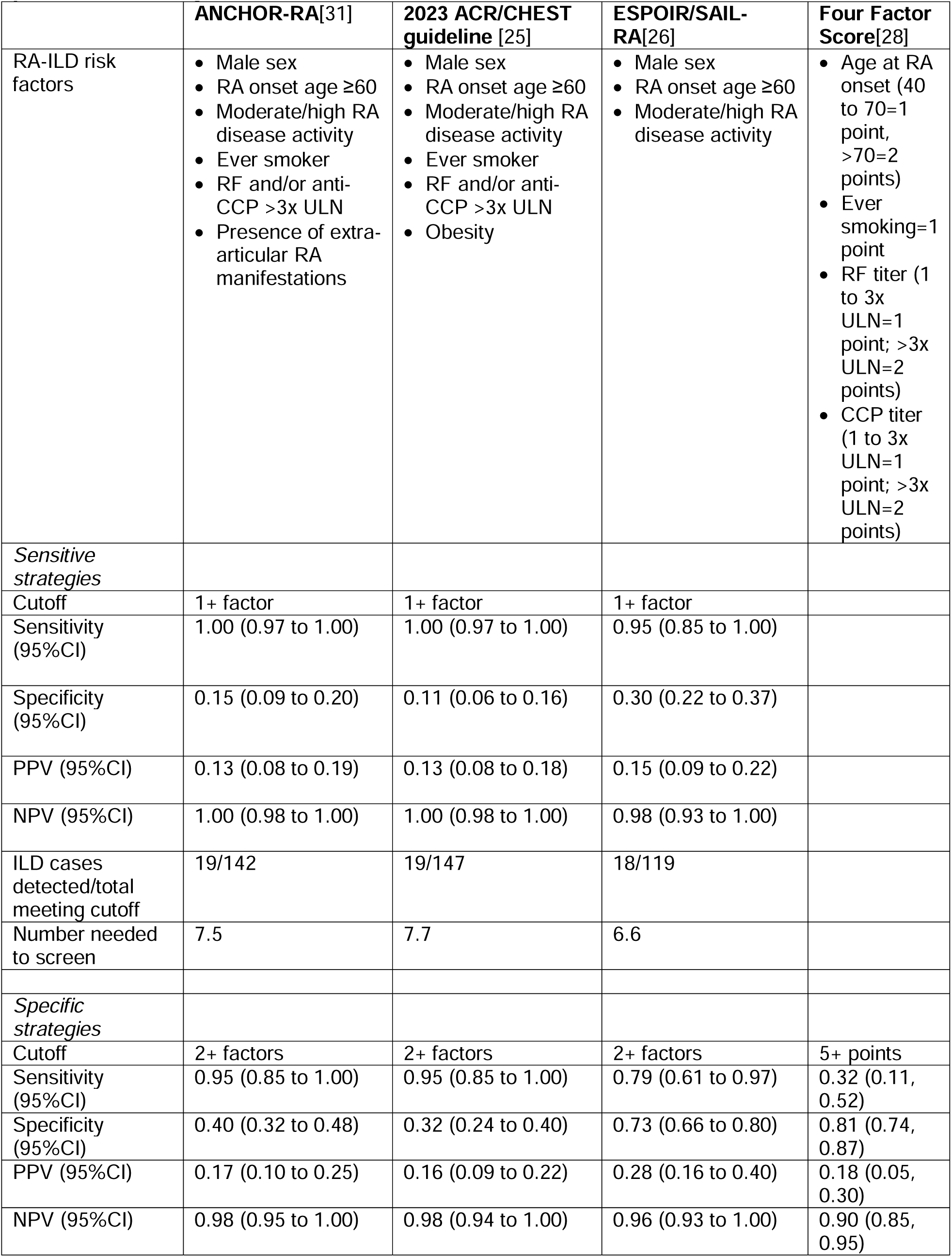
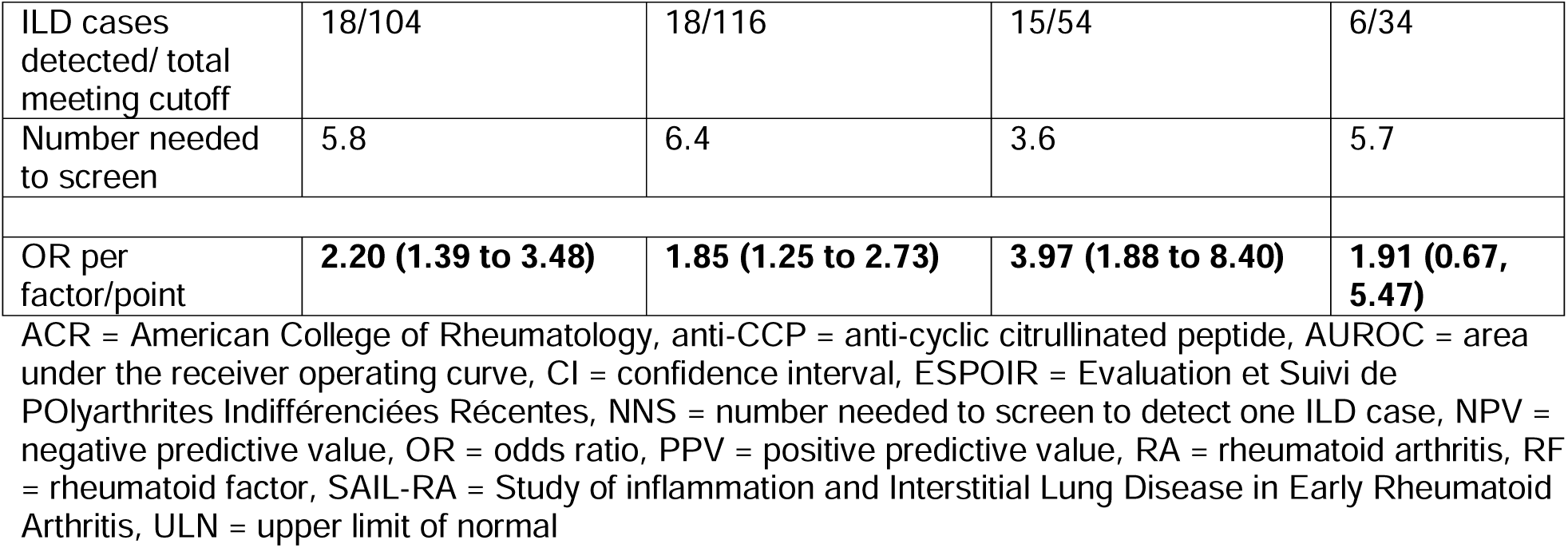
Performance of proposed strategies to screen for interstitial lung disease among patients with early RA in SAIL-RA.

We also reported the NNS to detect one patient with ILD for each set of criteria using a cutoff of one or two criteria. The NNS provides the number of early RA patients in our cohort who would need to be screened to detect one RA-ILD case. At a cutoff of two or more criteria, the NNS was 5.8 for ANCHOR-RA criteria, 5.8 for ACR/CHEST criteria, and 3.6 for ESPOIR/SAIL-RA criteria. The NNS using the 5-point cutoff of the Four Factor Score was 5.7. Area under the receiver operating curves (AUROC) for each set of criteria using all components are presented in **Figure 2**. Using all proposed factors in each strategy, the AUROC was 0.81 for ANCHOR-RA criteria, 0.80 for ESPOIR/SAIL-RA criteria, 0.81 for ACR/CHEST criteria, and 0.77 for the Four Factor Score.

**Figure 2:**
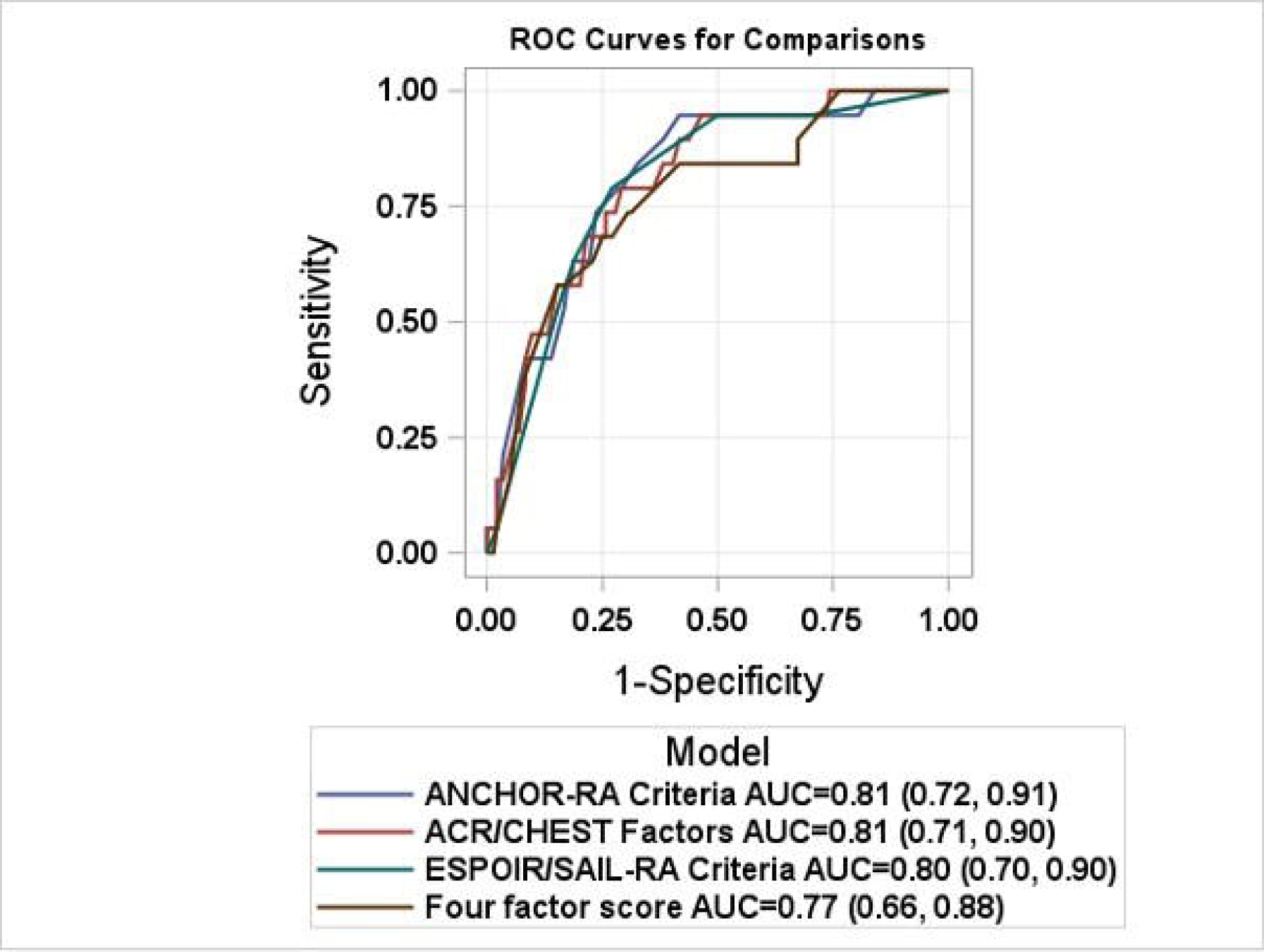
Area under the receiver operating curves for proposed RA-ILD screening strategies. ACR = American College of Rheumatology, ESPOIR = Evaluation et Suivi de POlyarthrites Indifférenciées Récentes, SAIL-RA = Study of inflammatory Arthritis and Interstitial Lung Disease in Early Rheumatoid Arthritis, ROC = receiver operating curve

After limiting to patients with MRC dyspnea score <3, we saw similar but slightly attenuated results (**Supplemental Table S5**). Using the cutoff of two or more criteria or 5 or more points for the Fourth Factor Score, the NNS was 4.1 for ESPOIR/SAIL-RA criteria, 5.5 for the Four Factor Score, 6.9 for ANCHOR-RA criteria, and 7.6 for the ACR/CHEST criteria.

## DISCUSSION

In this prospective, multicenter study, the prevalence of ILD on universal HRCT testing with independent expert thoracic radiologist assessment in early RA was 11%. Moderate or high RA disease activity during the early RA period was associated with 7-fold higher odds of ILD and >4-fold higher odds of any parenchymal lung disease. These findings suggest that disease activity in the early RA period may play a pivotal role in the development of RA-ILD and other RA-related lung diseases as well as serve as a highly informative risk factor for RA-ILD screening. We also present the first external validation, and application to an early RA population, for several RA-ILD screening strategies, including the 2023 ACR/CHEST screening guideline. These demonstrated high sensitivity and required 4-8 patients to be screened to detect an ILD case. The simplest model consisted of age at RA onset ≥60 years, male sex, and active RA, suggesting these three factors, which are routinely available at the point of care, may be suitable to risk stratify for ILD risk in early RA.

Our study adds to prior literature investigating lung disease in the early RA period and risk factors for RA-ILD. One prior study published in 1997 assessed patients with <2 years of RA joint disease and found that 58% had abnormalities consistent with ILD on an assessment that included HRCT, bronchoalveolar lavage, and nuclear scanning[16]. One study from Sweden investigated 105 RA patients with early, untreated RA using HRCT. This study found that 57 participants (54%) had some form of parenchymal lung abnormalities[14]. However, parenchymal lung abnormalities in this study were broadly defined and included nodules and did not specifically examine interstitial lung abnormalities. A total of 12 study participants (11%) had fibrosis, similar to the prevalence that we reported for RA-ILD in our study. Another study performed in Japan investigated 65 patients with RA of less than 1 year duration and found that 13.8% had classical ILD pattern on HRCT[19]. Notably, the prevalence of classical ILD was 26% in longstanding RA participants in the same study. While the prevalence of ILD in RA has varied substantially and been reported as high as 40-50%, our findings suggest that in the early RA period in the modern treatment era, the prevalence of clinical and subclinical ILD is more likely between 10 and 15%.

The study findings also recapitulate several known RA-ILD risk factors and emphasize their importance in the early RA period. We found moderate/high RA disease activity was strongly associated with RA-ILD in our cohort of early RA patients. RA disease activity has been previously noted to be a risk factor for incident RA-ILD in cohorts of established RA patients[45,46]. Notably, one study performed in a prospective cohort of RA patients (mean RA duration of 9 years) noted moderate or high disease activity measured by DAS28 was associated with three-fold increased risk of developing RA-ILD[45]. We also found older age at RA diagnosis is a risk factor for RA-ILD, which is consistent with observations from several prior studies[11,46–49]. Other prior studies have established male sex[11,46–49]and smoking[11,46,47] as key risk factors for RA-ILD. In our study, both risk factors were enriched in the RA-ILD patients and showed a trend towards association with RA-ILD but were not statistically significantly associated in multivariable models. However, our study included only 38 male patients and the prevalence of current or former smokers was only 38%, so we may have been underpowered to detect statistical associations for these factors.

There has been significant interest in the early RA period as a “window of opportunity” where timely use of effective immunosuppressive therapies can reduce the progression and severity of the subsequent articular RA disease course[21,22]. One study performed in two large European cohorts examined the association of symptom duration with favorable outcomes like subsequent remission with or without DMARD treatment[22]. The authors observed a nonlinear relationship between shorter symptom duration and favorable outcomes, suggesting that early treatment may be most beneficial before 15-19 weeks of symptoms[22]. Further mechanistic support for the importance of the early RA period comes from research demonstrating distinct cytokine profiles in synovial fluid of early RA patients compared to those with established RA[50]. In our study, we noted a strong association between disease activity and RA-ILD in the early RA period, suggesting early and aggressive treatment of RA disease activity may also impact the natural history of RA-ILD prior to symptom onset and clinical detection. Future treatment studies addressing the roles of disease activity and specific DMARDs in RA-ILD risk are needed.

Our study has several strengths to consider. First, to our knowledge, this is the largest lung screening study performed in early RA patients and was performed in multiple centers across the US. Second, we comprehensively assessed for evidence of RA-ILD in all participants using HRCT and also performed detailed assessment of medical history, pulmonary function, and disease activity status. Third, we were able to externally validate the performance of multiple proposed RA-ILD screening strategies in early RA patients, including the recently published 2023 ACR/CHEST screening guideline[25]. Early RA is a timepoint in the disease course that would likely be of significant interest for future screening programs and ultimately for treatment strategies aimed at reducing the burden of ILD among RA patients.

Our study also has potential limitations to consider. First, consistent with typical RA populations, most patients in SAIL-RA were female, which may have limited our power to detect differences between risk factor profiles in males and females. Second, although SAIL-RA was a multicenter study conducted at five sites in the United States, all the sites were academic medical centers, and this may limit the generalizability of the findings. Additionally, although the characteristics of the SAIL-RA cohort -- including sex, seropositive status, and medication use – mirror other RA cohorts, it remains possible that patients willing to participate in lung screening studies have unmeasured differences when compared to the general RA population. Third, visual assessment of HRCT for evidence of clinical and subclinical ILD is labor intensive and requires significant expertise from experienced thoracic radiologists. Future studies to investigate the use of automated quantitative CT imaging analysis and to augment visual scoring are needed. Furthermore, whether the performance of the RA-ILD screening strategies assessed in this study will perform similarly in routine clinical care remains to be determined.

Fourth, we did not have access to genetic data for SAIL-RA participants, including the *MUC5B* promoter variant, which is an established RA-ILD genetic risk factor. However, this is also not currently available in routine clinical care. Further studies to investigate genetic risk factors in early RA-ILD are needed. Finally, we performed cross-sectional analyses of baseline visits.

Follow-up in SAIL-RA is ongoing to investigate incidence and progression of RA-ILD after baseline. Future studies are needed to investigate the impact of early detection and treatment on RA-ILD outcomes.

In conclusion, we prospectively investigated the prevalence of RA-ILD and other parenchymal lung diseases in a cohort of patients with early RA. The prevalence of RA-ILD was 11% among patients in early RA, and moderate or high RA disease activity was associated with 6-fold higher odds of ILD. Finally, several RA-ILD screening strategies were assessed in patients with early RA. The simplest screening strategy incorporating older age, male sex, and active RA may be a useful tool available to providers at the point of care to inform ILD screening in the early RA period.

## Data availability statement

Data are available upon reasonable request and with appropriate institutional review board approval.

## Acknowledgements

We would like to acknowledge the SAIL-RA participants and study staff for their valuable contributions to this study.

## REFERENCES

1 Bongartz T, Nannini C, Medina-Velasquez YF, et al. Incidence and mortality of interstitial lung disease in rheumatoid arthritis - A population-based study. Arthritis Rheum. 2010;62:1583–91. doi: 10.1002/art.27405

2 Hyldgaard C, Hilberg O, Pedersen AB, et al. A population-based cohort study of rheumatoid arthritis-associated interstitial lung disease: Comorbidity and mortality. Ann Rheum Dis. 2017;76:1700–6. doi: 10.1136/annrheumdis-2017-211138

3 Kim D, Cho SK, Choi CB, et al. Impact of interstitial lung disease on mortality of patients with rheumatoid arthritis. Rheumatol Int. 2017;37:1735–45. doi: 10.1007/s00296-017-3781-7

4 Duarte AC, Porter JC, Leandro MJ. The lung in a cohort of rheumatoid arthritis patients-an overview of different types of involvement and treatment. Rheumatology (Oxford). 2019;58:2031–8. doi: 10.1093/rheumatology/kez177

5 Wang D, Zhang J, Lau J, et al. Mechanisms of lung disease development in rheumatoid arthritis. Nat Rev Rheumatol. 2019;15:581–96. doi: 10.1038/s41584-019-0275-x

6 Raimundo K, Solomon JJ, Olson AL, et al. Rheumatoid Arthritis-Interstitial Lung Disease in the United States: Prevalence, Incidence, and Healthcare Costs and Mortality. J Rheumatol. 2019;46:360–9. doi: 10.3899/jrheum.171315

7 Samhouri BF, Vassallo R, Achenbach SJ, et al. Incidence, Risk Factors, and Mortality of Clinical and Subclinical Rheumatoid Arthritis-Associated Interstitial Lung Disease: A Population-Based Cohort. Arthritis Care Res (Hoboken). 2022;74:2042–9. doi: 10.1002/acr.24856

8 Doyle TJ, Dellaripa PF, Batra K, et al. Functional impact of a spectrum of interstitial lung abnormalities in rheumatoid arthritis. Chest. 2014;146:41–50. doi: 10.1378/chest.13-1394

9 McDermott GC, Doyle TJ, Sparks JA. Interstitial lung disease throughout the rheumatoid arthritis disease course. Curr Opin Rheumatol. 2021;33:284–91. doi: 10.1097/BOR.0000000000000787

10 Mohning MP, Amigues I, Demoruelle MK, et al. Duration of rheumatoid arthritis and the risk of developing interstitial lung disease. ERJ Open Res. 2021;7:00633–2020. doi: 10.1183/23120541.00633-2020

11 Kelly CA, Saravanan V, Nisar M, et al. Rheumatoid arthritis-related interstitial lung disease: Associations, prognostic factors and physiological and radiological characteristics-a large multicentre UK study. Rheumatology (United Kingdom). 2014;53:1676–82. doi: 10.1093/rheumatology/keu165

12 Zhang Y, Li H, Wu N, et al. Retrospective study of the clinical characteristics and risk factors of rheumatoid arthritis-associated interstitial lung disease. Clin Rheumatol. 2017;36:817–23. doi: 10.1007/s10067-017-3561-5

13 Chen J, Shi Y, Wang X, et al. Asymptomatic preclinical rheumatoid arthritis-associated interstitial lung disease. Clin Dev Immunol. 2013;2013:10–5. doi: 10.1155/2013/406927

14 Reynisdottir G, Karimi R, Joshua V, et al. Structural changes and antibody enrichment in the lungs are early features of anti-citrullinated protein antibody-positive rheumatoid arthritis. Arthritis and Rheumatology. 2014;66:31–9. doi: 10.1002/art.38201

15 Doyle JJ, Eliasson AH, Argyros GJ, et al. Prevalence of pulmonary disorders in patients with newly diagnosed rheumatoid arthritis. Clin Rheumatol. 2000;19:217–21. doi: 10.1007/s100670050160

16 Gabbay E, Tarala R, Will R, et al. Interstitial lung disease in recent onset rheumatoid arthritis. Am J Respir Crit Care Med. 1997;156:528–35. doi: 10.1164/ajrccm.156.2.9609016

17 Habib HM, Eisa AA, Arafat WR, et al. Pulmonary involvement in early rheumatoid arthritis patients. Clin Rheumatol. 2011;30:217–21. doi: 10.1007/s10067-010-1492-5

18 Dong H, Julien PJ, Demoruelle MK, et al. Interstitial lung abnormalities in patients with early rheumatoid arthritis: A pilot study evaluating prevalence and progression. Eur J Rheumatol. 2019;6:193–8. doi: 10.5152/eurjrheum.2019.19044

19 Mori S, Cho I, Koga Y, et al. Comparison of pulmonary abnormalities on high-resolution computed tomography in patients with early versus longstanding rheumatoid arthritis. Journal of Rheumatology. 2008;35:1513–21. doi: 10.1016/s0098-1672(09)79355-5

20 Holers VM, Demoruelle MK, Kuhn KA, et al. Rheumatoid arthritis and the mucosal origins hypothesis: protection turns to destruction. Nat Rev Rheumatol. 2018;14:542–57. doi: 10.1038/s41584-018-0070-0

21 Boers M. Understanding the window of opportunity concept in early rheumatoid arthritis. Arthritis Rheum. 2003;48:1771–4. doi: 10.1002/art.11156

22 van Nies JAB, Tsonaka R, Gaujoux-Viala C, et al. Evaluating relationships between symptom duration and persistence of rheumatoid arthritis: does a window of opportunity exist? Results on the Leiden early arthritis clinic and ESPOIR cohorts. Ann Rheum Dis. 2015;74:806–12. doi: 10.1136/annrheumdis-2014-206047

23 Burgers LE, Raza K, van der Helm-van Mil AH. Window of opportunity in rheumatoid arthritis - definitions and supporting evidence: from old to new perspectives. RMD Open. 2019;5:e000870. doi: 10.1136/rmdopen-2018-000870

24 Matteson EL, Matucci-Cerinic M, Kreuter M, et al. Patient-level factors predictive of interstitial lung disease in rheumatoid arthritis: a systematic review. RMD Open. 2023;9. doi: 10.1136/rmdopen-2023-003059

25 Johnson SR, Bernstein EJ, Bolster MB, et al. 2023 American College of Rheumatology (ACR)/American College of Chest Physicians (CHEST) Guideline for the Screening and Monitoring of Interstitial Lung Disease in People with Systemic Autoimmune Rheumatic Diseases. Arthritis Rheumatol. 2024;76:1201–13. doi: 10.1002/art.42860

26 Juge P-A, Granger B, Debray M-P, et al. A Risk Score to Detect Subclinical Rheumatoid Arthritis-Associated Interstitial Lung Disease. Arthritis Rheumatol. 2022;74:1755–65. doi: 10.1002/art.42162

27 Juge P-A, Lee JS, Ebstein E, et al. MUC5B Promoter Variant and Rheumatoid Arthritis with Interstitial Lung Disease . New England Journal of Medicine. 2018;379:2209–19. doi: 10.1056/nejmoa1801562

28 Koduri GM, Podlasek A, Pattapola S, et al. Four-factor risk score for the prediction of interstitial lung disease in rheumatoid arthritis. Rheumatol Int. 2023;43:1515–23.

29 Narváez J, Aburto M, Seoane-Mato D, et al. Screening criteria for interstitial lung disease associated to rheumatoid arthritis: Expert proposal based on Delphi methodology. Reumatol Clin. 2023;19:74–81. doi: 10.1016/j.reumae.2021.12.003

30 Paulin F, Doyle TJ, Mercado JF, et al. Development of a risk indicator score for the identification of interstitial lung disease in patients with rheumatoid arthritis. Reumatol Clin. 2021;17:207–11. doi: 10.1016/j.reuma.2019.05.007

31 Sparks JA, Dieudé P, Hoffmann-Vold A-M, et al. Design of ANCHOR-RA: a multi-national cross-sectional study on screening for interstitial lung disease in patients with rheumatoid arthritis. BMC Rheumatol. 2024;8:19. doi: 10.1186/s41927-024-00389-4

32 Aletaha D, Neogi T, Silman AJ, et al. 2010 Rheumatoid arthritis classification criteria: An American College of Rheumatology/European League Against Rheumatism collaborative initiative. Arthritis Rheum. 2010;62:2569–81. doi: 10.1002/art.27584

33 Mahler DA, Wells CK. Evaluation of clinical methods for rating dyspnea. Chest. 1988;93:580–6. doi: 10.1378/chest.93.3.580

34 van der Heijde DM, van ’t Hof MA, van Riel PL, et al. Judging disease activity in clinical practice in rheumatoid arthritis: first step in the development of a disease activity score. Ann Rheum Dis. 1990;49:916–20. doi: 10.1136/ard.49.11.916

35 Aletaha D, Nell VPK, Stamm T, et al. Acute phase reactants add little to composite disease activity indices for rheumatoid arthritis: validation of a clinical activity score. Arthritis Res Ther. 2005;7:R796–806. doi: 10.1186/ar1740

36 Hatabu H, Hunninghake GM, Richeldi L, et al. Interstitial lung abnormalities detected incidentally on CT: a Position Paper from the Fleischner Society. Lancet Respir Med. 2020;8:726–37. doi: 10.1016/S2213-2600(20)30168-5

37 Tashkin DP, Roth MD, Clements PJ, et al. Mycophenolate mofetil versus oral cyclophosphamide in scleroderma-related interstitial lung disease (SLS II): a randomised controlled, double-blind, parallel group trial. Lancet Respir Med. 2016;4:708–19. doi: 10.1016/S2213-2600(16)30152-7

38 Raghu G, Remy-Jardin M, Myers JL, et al. Diagnosis of idiopathic pulmonary fibrosis An Official ATS/ERS/JRS/ALAT Clinical practice guideline. Am J Respir Crit Care Med. 2018;198:e44–68. doi: 10.1164/rccm.201807-1255ST

39 Kligerman SJ, Groshong S, Brown KK, et al. Nonspecific interstitial pneumonia: radiologic, clinical, and pathologic considerations. Radiographics. 2009;29:73–87. doi: 10.1148/rg.291085096

40 Lynch DA, Sverzellati N, Travis WD, et al. Diagnostic criteria for idiopathic pulmonary fibrosis: a Fleischner Society White Paper. Lancet Respir Med. 2018;6:138–53.

41 Rasch EK, Hirsch R, Paulose-Ram R, et al. Prevalence of rheumatoid arthritis in persons 60 years of age and older in the United States: effect of different methods of case classification. Arthritis Rheum. 2003;48:917–26. doi: 10.1002/art.10897

42 Li X, Cesta A, Movahedi M, et al. Late-onset rheumatoid arthritis has a similar time to remission as younger-onset rheumatoid arthritis: results from the Ontario Best Practices Research Initiative. Arthritis Res Ther. 2022;24:255. doi: 10.1186/s13075-022-02952-1

43 Clopper CJ, Pearson ES. The Use of Confidence or Fiducial Limits Illustrated in the Case of the Binomial. Biometrika. 1934;26:404–13. doi: 10.1093/biomet/26.4.404

44 Sparks JA, Doyle TJ, He X, et al. Incidence and predictors of dyspnea on exertion in a prospective cohort of patients with rheumatoid arthritis. ACR Open Rheumatol. 2019;1:4–15. doi: 10.1002/acr2.1001

45 Sparks JA, He X, Huang J, et al. Rheumatoid Arthritis Disease Activity Predicting Incident Clinically Apparent Rheumatoid Arthritis–Associated Interstitial Lung Disease: A Prospective Cohort Study. Arthritis and Rheumatology. 2019;71:1472–82. doi: 10.1002/art.40904

46 Restrepo JF, del Rincón I, Battafarano DF, et al. Clinical and laboratory factors associated with interstitial lung disease in rheumatoid arthritis. Clin Rheumatol. 2015;34:1529–36. doi: 10.1007/s10067-015-3025-8

47 Doyle TJ, Patel AS, Hatabu H, et al. Detection of rheumatoid arthritis-interstitial lung disease is enhanced by serum biomarkers. Am J Respir Crit Care Med. 2015;191:1403–12. doi: 10.1164/rccm.201411-1950OC

48 Saag KG, Kolluri S, Koehnke RK, et al. Rheumatoid arthritis lung disease: Determinants of radiographic and physiologic abnormalities. Arthritis Rheum. 1996;39:1711–9. doi: 10.1002/art.1780391014

49 Kiely P, Busby AD, Nikiphorou E, et al. Is incident rheumatoid arthritis interstitial lung disease associated with methotrexate treatment? Results from a multivariate analysis in the ERAS and ERAN inception cohorts. BMJ Open. 2019;9. doi: 10.1136/bmjopen-2018-028466

50 Raza K, Falciani F, Curnow SJ, et al. Early rheumatoid arthritis is characterized by a distinct and transient synovial fluid cytokine profile of T cell and stromal cell origin. Arthritis Res Ther. 2005;7:R784–95. doi: 10.1186/ar1733

