## Supplement for "Risk factors for interstitial lung disease in early rheumatoid arthritis and external validation of screening strategies: Baseline results of SAIL-RA"

**Supplemental Table S1: Baseline characteristics of SAIL-RA participants by presence or absence of parenchymal lung disease**

| **Characteristic** | **Overall sample**  **(n=172)** | **ILD, emphysema, or bronchiectasis (n=28)** | **No parenchymal lung disease**  **(n=144)** |
| --- | --- | --- | --- |
| **Demographics** |  |  |  |
| Age, years (Mean, SD) | 55.3 (13.7) | 64.3 (12.7) | 53.5 (13.3) |
| Sex at birth |  |  |  |
| Male | 44 (25.6%) | 12 (42.9%) | 32 (22.2%) |
| Female | 128 (74.4%) | 16 (57.1%) | 112 (77.8%) |
| Race |  |  |  |
| Asian | 8 (4.7%) | 0 (0.0%) | 8 (5.6%) |
| Black | 10 (5.8%) | 2 (7.1%) | 8 (5.6%) |
| White | 144 (83.7%) | 25 (89.3%) | 119 (82.6%) |
| Other | 10 (5.8%) | 1 (3.6%) | 9 (6.3%) |
| Hispanic | 15 (8.7%) | 3 (10.7%) | 12 (8.3%) |
| **Lifestyle** |  |  |  |
| Smoking status |  |  |  |
| Never | 106 (61.6%) | 12 (42.9%) | 94 (65.3%) |
| Past | 55 (32.0%) | 13 (46.4%) | 42 (29.2%) |
| Current | 11 (6.4%) | 3 (10.7%) | 8 (5.6%) |
| Smoking pack-years (median, IQR) | 0 (0, 9.5) | 4.5 (0, 20.5) | 0 (0, 4.23) |
| BMI, median (IQR) | 28.1 (24.5, 33.0) | 26.2 (24.4, 30.9) | 28.2 (24.5, 33.9) |
| **RA characteristics** |  |  |  |
| RA diagnosis duration, years (median, IQR) | 0.79 (0.36, 1.60) | 0.81 (0.28, 1.62) | 0.78 (0.37, 1.58) |
| RA symptom duration, years (median, IQR) | 1.62 (0.86, 2.47) | 1.8 (0.78, 2.31) | 1.57 (0.88, 2.50) |
| DAS28-ESR (median, IQR) | 3.08 (1.90, 4.28) | 3.75 (3.11, 4.39) | 2.8 (1.75, 4.27) |
| TJC28 (median, IQR) | 1 (0, 5) | 2 (1, 5) | 0.5 (0, 4) |
| SJC28 (median, IQR) | 1 (0, 3) | 1.5 (0, 4) | 0.5 (0, 3) |
| GH (median, IQR) | 20 (10, 50) | 30 (10, 60) | 20 (10, 40) |
| ESR (median, IQR) | 15 (6, 32.5) | 19 (10, 37) | 14 (6, 29) |
| Disease activity |  |  |  |
| Remission/low | 93 (54.1%) | 9 (32.1%) | 84 (58.3%) |
| Moderate/high | 79 (45.9%) | 19 (67.9%) | 60 (41.7%) |
| Seropositive | 140/171 (81.9%) | 22/28 (78.6%) | 118/143 (82.5%) |
| Anti-CCP positive | 126/171 (73.7%) | 20/28 (71.4%) | 106/143 (74.1%) |
| Anti-CCP 1-3x ULN | 18/168 (10.7%) | 2/27 (7.4%) | 16/141 (11.4%) |
| Anti-CCP >3x ULN | 73/168 (43.5%) | 14/27 (51.9%) | 59/141 (41.8%) |
| RF positive | 116/170 (68.2%) | 21/28 (75.0%) | 95/142 (66.9%) |
| RF titer 1-3x ULN | 21/167 (12.6%) | 3/27 (11.1%) | 18/140 (12.9%) |
| RF titer >3x ULN | 40/167 (24.0%) | 12/27 (44.4%) | 28/140 (20.0%) |
| Presence of extra-articular RA manifestations | 9 (5.2%) | 1 (3.6%) | 8 (5.6%) |
| Current medications |  |  |  |
| Glucocorticoids | 111 (64.5%) | 17 (60.7%) | 94 (65.3%) |
| Methotrexate | 65 (37.8%) | 9 (32.1%) | 56 (38.9%) |
| Other csDMARDs | 50 (29.1%) | 7 (25.0%) | 43 (29.9%) |
| TNF inhibitors | 33 (19.2%) | 6 (21.4%) | 27 (18.8%) |
| **PFTs** |  |  |  |
| FEV_1_/FVC (median, IQR) | 0.79 (0.74, 0.82) | 0.77 (0.71, 0.8) | 0.79 (0.74, 0.83) |
| FEV_1_% (median, IQR) | 96.1 (83.0, 106) | 93.8 (74.7, 105) | 96.6 (84.8, 106) |
| FVC% (median, IQR) | 96.3 (87.0, 107) | 93.2 (78.4, 110) | 97 (87, 107) |
| DLCO% (median, IQR) | 88.1 (76.6, 99.0) | 76.5 (69.0, 85.0) | 91 (79.0, 100) |
| FVC%<80% | 25/162 (15.4%) | 9/26 (34.6%) | 16/136 (11.8%) |
| FEV_1_/FVC<70% | 22/163 (13.5%) | 5/26 (19.2%) | 17/137 (12.4%) |
| DLCO%<75% | 31/146 (21.2%) | 9/22 (40.9%) | 22/124 (17.7%) |
| **MRC dyspnea score, continuous (median)** | 0 (0, 1) | 0 (0, 2) | 0 (0, 1) |
| MRC dyspnea score >=3 | 12/172 (7%) | 6/28 (21.4%) | 6/144 (4.2%) |

Anti-CCP = anti-cyclic citrullinated peptide, BMI = body mass index, csDMARD = conventional synthetic disease modifying antirheumatic drug, DAS28-ESR = disease activity score with 28 joints with ESR, DLCO = diffusion capacity of carbon monoxide, ESR = erythrocyte sedimentation rate, FEV1 = forced expiratory volume in 1 second, FVC = forced vital capacity, GH = global health, MRC = Modified Medical Research Council, SD = standard deviation, SJC = swollen joint count, TJC = tender joint count, TNF = tumor necrosis factor, RF = rheumatoid factor, ULN = upper limit of normal

**Supplemental Table S2: Semiquantitative HRCT Chest scoring and pulmonary function testing in RA-ILD cases (n=19)**

| **Age** | **Sex** | **Radiologic Subtype** | **Semiquantitative % Involvement** |  | **FEV_1_ % predicted** | **FVC % predicted** | **DLCO % predicted** | **FEV_1_/FVC** |
| --- | --- | --- | --- | --- | --- | --- | --- | --- |
| 70-79 | M | Probable UIP | >30% |  | 105 | 90 | 16 | 0.88 |
| 70-79 | F | NSIP | >30% |  | 128 | 119 | 52 | 0.82 |
| 60-69 | F | Indeterminate for UIP (UIP vs. NSIP) | 10-30% |  | 94.6 | 93.9 | 54.1 | 0.78 |
| 50-59 | F | Unclassified | 10-30% |  | 114 | 110 | 85 | 0.81 |
| 60-69 | F | Probable UIP | 10-30% |  | 93 | 91 | 51 | 0.80 |
| 50-59 | F | HP | 10-30% |  | 76.6 | 78.8 | 75.5 | 0.82 |
| 70-79 | F | NSIP | 10-30% |  | 120 | 120 | 78 | 0.78 |
| 60-69 | F | NSIP | 0-10% |  | 102.9 | 99.5 | 79 | 0.80 |
| 70-79 | M | Probable UIP | 0-10% |  | 88.7 | 102.5 | 72.4 | 0.65 |
| 70-79 | F | Unclassified | 0-10% |  |  |  |  |  |
| 60-69 | F | Probable UIP | 0-10% |  | 74 | 71 | 84 | 0.81 |
| 60-69 | F | NSIP | 0-10% |  | 74.7 | 78.4 | 84.6 | 0.76 |
| 60-69 | F | Unclassified | 0-10% |  | 71 | 76.9 | 75.5 | 0.71 |
| 40-49 | M | Unclassified | 0-10% |  | 83 | 83 |  | 0.79 |
| 70-79 | M | Unclassified | 0-10% |  | 51 | 55 |  | 0.67 |
| 50-59 | F | NSIP | 0-10% |  | 78 | 79 |  | 0.76 |
| 80-89 | M | Probable UIP | 0-10% |  | 125 | 128 | 89 | 0.72 |
| 50-59 | M | Unclassified | 0-10% |  |  |  |  |  |
| 30-39 | F | Unclassified | 0-10% |  | 99 | 104 |  | 0.79 |

DLCO = diffusion capacity of carbon monoxide, FEV1 = forced expiratory volume in one second, FVC = forced vital capacity, HP = hypersensitivity pneumonitis, NSIP = nonspecific interstitial pneumonia, UIP = usual interstitial pneumonia

**Supplemental Table S3: Odds ratios for RA-ILD in early RA by baseline characteristics, additionally adjusted for RF level and smoking status**

| **Characteristic** | **RA-ILD cases/denominator in stratum (%)** | **Unadjusted OR (95%CI)** | **Multivariable* OR (95%CI)** |
| --- | --- | --- | --- |
| **Demographics** |  |  |  |
| Age at RA diagnosis |  |  |  |
| <60 years | 6/99 (6.1%) | 1.0 (Ref) | 1.0 (Ref) |
| ≥60 years | 13/64 (20.3%) | **3.95 (1.42, 11.02)** | **3.30 (1.10, 9.89)** |
| Sex at birth |  |  |  |
| Male | 6/38 (15.8%) | 1.62 (0.57, 4.59) | 2.49 (0.73, 8.55) |
| Female | 13/125 (10.4%) | 1.0 (Ref) | 1.0 (Ref) |
| **Lifestyle** |  |  |  |
| Smoking status |  |  | - |
| Never | 10/104 (9.6%) | 1.0 (Ref) | 1.0 (Ref) |
| Ever | 9/59 (15.3%) | 1.69 (0.65, 4.44) | 1.33 (0.45, 3.96) |
| Pack-years (per unit) | 19/163 (11.7%) | 1.00 (0.99, 1.01) | - |
| Body mass index |  |  |  |
| Underweight/normal | 4/43 (9.3%) | 1.0 (Ref) | - |
| Overweight | 8/58 (13.8%) | 1.56 (0.44, 5.56) | - |
| Obese | 7/62 (11.3%) | 1.24 (0.34, 4.53) | - |
| **RA characteristics** |  |  |  |
| Anti-CCP level |  |  |  |
| Negative | 8/74 (10.8%) | 1.0 (Ref) | - |
| >1 to 3x ULN | 1/17 (5.9%) | 0.52 (0.06, 4.42) | - |
| >3x ULN | 9/68 (13.2%) | 1.26 (0.46, 3.47) | - |
| RF level |  |  |  |
| Negative | 8/102 (7.8%) | 1.0 (Ref) | 1.0 (Ref) |
| >1 to 3x ULN | 3/21 (14.3%) | 1.96 (0.47, 8.10) | 1.40 (0.31, 6.29) |
| >3x ULN | 7/35 (20.0%) | 2.94 (0.98, 8.81) | 1.63 (0.48, 5.47) |
| Extra-articular RA manifestations |  |  |  |
| No | 18/154 (11.7%) | 1.0 (Ref) | - |
| Yes | 1/9 (11.1%) | 0.94 (0.11, 8.00) | - |
| RA disease activity |  |  |  |
| Remission/low | 4/88 (4.6%) | 1.0 (Ref) | 1.0 (Ref) |
| Moderate/high | 15/75 (20.0%) | **5.25 (1.66, 16.6)** | **5.39 (1.44, 20.19)** |

* adjusted for age at RA diagnosis, sex, and RA disease activity

For 12 patients with missing ESR values, disease activity category was determined using CDAI.

Anti-CCP = anti-cyclic citrullinated peptide, CI = confidence interval, ILD = interstitial lung disease, OR = odds ratio, RA = rheumatoid arthritis, RF = rheumatoid factor, ULN = upper limit of normal

**Supplemental Table S4: Multivariable odds ratios for ILD in early RA, stratified by sex at birth.**

Females (n=125)

| **Characteristic** | **ILD cases/denominator in stratum** | **Unadjusted OR (95%CI)** | **Multivariable OR (95%CI)** |
| --- | --- | --- | --- |
| **Demographics** |  |  |  |
| Age at RA diagnosis |  |  |  |
| <60 years | 4/80 (5.0%) | 1.0 (Ref) | 1.0 (Ref) |
| ≥60 years | 9/45 (20.0%) | **4.75 (1.37, 16.5)** | **3.87 (1.07, 14.0)** |
| **Lifestyle** |  |  |  |
| Smoking status |  |  | - |
| Never | 7/82 (8.5%) | 1.0 (Ref) | - |
| Ever | 6/43 (14.0%) | 1.74 (0.55, 5.54) | - |
| Pack-years (per unit) | 13/125 (10.4%) | 1.00 (0.99, 1.01) | - |
| **RA disease activity** |  |  |  |
| Remission/low | 1/61 (1.6%) | 1.0 (Ref) | 1.0 (Ref) |
| Moderate/high | 12/64 (18.8%) | **13.8 (1.74, 110)** | **11.9 (1.47, 95.9)** |

Males (n=38)

| **Characteristic** | **ILD cases/denominator in stratum** | **Unadjusted OR (95%CI)** | **Multivariable OR (95%CI)** |
| --- | --- | --- | --- |
| **Demographics** |  |  |  |
| Age at RA diagnosis |  |  |  |
| <60 years | 2/19 (10.5%) | 1.0 (Ref) | 1.0 (Ref) |
| ≥60 years | 4/19 (21.1%) | 2.27 (0.36, 14.2) | 3.08 (0.43, 22.1) |
| **Lifestyle** |  |  |  |
| Smoking status |  |  | - |
| Never | 3/22 (13.6%) | 1.0 (Ref) | - |
| Ever | 3/16 (18.8%) | 1.46 (0.25, 8.40) | - |
| Pack-years (per unit) | 6/38 (15.8%) | 1.00 (0.95, 1.05) | - |
| **RA disease activity** |  |  |  |
| Remission/low | 3/27 (11.1%) | 1.0 (Ref) | 1.0 (Ref) |
| Moderate/high | 3/11 (27.3%) | 3.00 (0.50, 18.0) | 3.92 (0.58, 26.6) |

CI = confidence interval, ILD = interstitial lung disease, OR = odds ratio, RA = rheumatoid arthritis

**Supplementary Table S5: Performance of proposed strategies to screen for interstitial lung disease among early RA participants with MRC dyspnea score <3 in SAIL-RA**

|  | **ANCHOR-RA**^31^ | **2023 ACR/CHEST guideline** ^25^ | **ESPOIR/SAIL-RA**^26^ | **Four Factor Score**^28^ |
| --- | --- | --- | --- | --- |
| RA-ILD risk factors | - Male sex - RA onset age ≥60 - Moderate/high RA disease activity - Ever smoker - RF and/or anti-CCP >3x ULN - Presence of extra-articular RA manifestations | - Male sex - RA onset age ≥60 - Moderate/high RA disease activity - Ever smoker - RF and/or anti-CCP >3x ULN - Obesity | - Male sex - RA onset age ≥60 - Moderate/high RA disease activity | - Age at RA onset (40 to 70=1 point, >70=2 points) - Ever smoking=1 point - RF titer (1 to 3x ULN=1 point; >3x ULN=2 points) - CCP titer (1 to 3x ULN=1 point; >3x ULN=2 points) |
| *Sensitive strategies* |  |  |  |  |
| Cutoff | 1+ factor | 1+ factor | 1+ factor |  |
| Sensitivity (95%CI) | 1.00 (0.97 to 1.00) | 1.00 (0.97 to 1.00) | 0.93 (0.81 to 1.00) |  |
| Specificity (95%CI) | 0.14 (0.09 to 0.20) | 0.12 (0.06 to 0.17) | 0.30 (0.22 to 0.37) |  |
| PPV (95%CI) | 0.11 (0.06 to 0.17) | 0.11 (0.06 to 0.16) | 0.13 (0.06 to 0.19) |  |
| NPV (95%CI) | 1.00 (0.98 to 1.00) | 1.00 (0.98 to 1.00) | 0.98 (0.93 to 1.00) |  |
| ILD cases detected/total meeting cutoff | 15/133 | 15/137 | 14/111 |  |
| Number needed to screen | 8.9 | 9.1 | 7.9 |  |
| *Specific strategies* |  |  |  |  |
| Cutoff | 2+ factors | 2+ factors | 2+ factors | 5+ points |
| Specificity (95%CI) | 0.40 (0.32, 0.48) | 0.33 (0.25, 0.40) | 0.73 (0.66, 0.81) | 0.80 (0.74, 0.87) |
| Sensitivity (95%CI) | 0.93 (0.81, 1.00) | 0.93 (0.81, 1.00) | 0.80 (0.60, 1.00) | 0.40 (0.15, 0.65) |
| NPV (95%CI) | 0.98 (0.95, 1.00) | 0.98 (0.94, 1.00) | 0.97 (0.94, 1.00) | 0.93 (0.88, 0.97) |
| PPV (95%CI) | 0.14 (0.07, 0.21) | 0.13 (0.07, 0.19) | 0.24 (0.12, 0.37) | 0.18 (0.05, 0.31) |
| ILD cases detected/ total meeting cutoff | 14/97 | 14/107 | 12/49 | 6/33 |
| Number needed to screen | 6.9 | 7.6 | 4.1 | 5.5 |
| OR per factor/point | **2.70 (1.53, 4.75)** | **2.19 (1.37, 3.50)** | **4.35 (1.83, 10.32)** | 2.74 (0.90, 8.36) |

ACR = American College of Rheumatology, anti-CCP = anti-cyclic citrullinated peptide, AUROC = area under the receiver operating curve, CI = confidence interval, ESPOIR = Evaluation et Suivi de POlyarthrites Indifférenciées Récentes, NNS = number needed to screen to detect one ILD case, NPV = negative predictive value, OR = odds ratio, PPV = positive predictive value, RA = rheumatoid arthritis, RF = rheumatoid factor, SAIL-RA = Study of inflammation and Interstitial Lung Disease in Early Rheumatoid Arthritis, ULN = upper limit of normal
